# Prevalence and risk factors for human papillomavirus infection among women living with HIV in Sierra Leone: A cross-sectional study

**DOI:** 10.64898/2026.11.11.26367873

**Authors:** Darlinda F. Jiba, Guang Yang, Tian Zhang, Lili Zhao, J. Andrew Dykens, Gibrilla F. Deen, Enanga S. Namanga, Matilda N. Kamara, Umu Barrie, Abu Bakarr Kallon, Daniel Sesay, Sattu Issa, Umu Sesay, Maima Amy Sam-Kpakra, Olukemi Adekanmbi, Babatunde Duduyemi, Sulaiman Lakoh

**Author notes:** Corresponding author: (DFJ). First co-authors (Darlinda F. Jiba, Guang Yang).

## Abstract

**Background:** Cervical cancer, a malignancy affecting the cervix, is the fourth most common cancer and cause of cancer-related mortality among women worldwide. This burden disproportionately affects low-resource regions, where approximately 90% of cases occur. HIV-positive women are at high risk of persistent Human Papillomavirus (HPV) infection and cervical cancer, but screening for this cancer is not yet integrated into HIV care in Sierra Leone. This study aimed to assess the prevalence and risk factors of high-risk HPV infection among women living with HIV (WLHIV) attending Connaught Hospital in Sierra Leone.

**Methods:** We conducted a cross-sectional study of women living with HIV aged 25-65 years attending the HIV clinic at Connaught Teaching Hospital in Freetown between September and December 2024. Endocervical swabs were collected for HPV DNA testing. Data was analyzed using R (v4.4.0). Logistic regression analysis was used to determine the independent risk factors for HPV positivity.

**Results:** A total of 191 WLHIV were enrolled. The mean age was 36.8 years; 44.5% were single, and 67% were employed. High-risk HPV DNA was detected in 60 (31.4%) (95% CI 25.3%-38.3%) participants, with 50% (30/60) having multiple serotypes. The most common genotype was HPV 52, followed by 16 and 51. Only 30% of HPV-positive women had genotypes 16 or 18, targeted by the bivalent vaccine. On multivariable analysis, former contraceptive use was independently associated with increased HPV positivity (aOR=4.86, 95% CI: 1.65-17.37, p = 0.0032).

**Conclusion:** The high prevalence of high-risk HPV infection and predominance of non 16/18 genotypes among WLHIV in Sierra Leone underscore the need to integrate HPV screening within HIV services and consider broader HPV vaccine coverage in Sierra Leone. Former contraceptive use is an independent predictor of HPV positivity, emphasizing the need for robust screening strategies in women using contraceptives.

## Background

Globally, cervical cancer is the fourth most common cancer and the fourth leading cause of cancer death in women, with an estimated 660,000 new cases and 350,000 deaths in 2022^1,2^. The global cervical cancer deaths are predicted to increase to 460,000 by 2040 without effective preventive interventions.^3^ In sub-Saharan Africa, it is the second most common cancer and the leading cause of cancer-related deaths among women^2^. About 95% of countries with the highest burden of cervical cancer in 2018 were in Sub-Saharan Africa^3^.

There are 500 new cases of cervical cancer in Sierra Leone each year, with an estimated 400 deaths. Although 1.85 million women aged 15 years and older are at risk of cervical cancer in Sierra Leone, the limited preventive services, high cost of pathology and surgical services, and the lack of radiotherapy impact service delivery.^4^

Human Papillomavirus (HPV) infection accounts for over 95% of all cervical cancers, highlighting that cervical cancer is a preventable malignancy that can be successfully treated if detected at an early stage. However, poor access to these facilities accounts for the late presentation and high mortality from cervical cancer \in Low- and Middle-Income Countries (LMICs).^1^

Cervical cancer is the most common cancer among women living with HIV worldwide.^2^ HIV increases the likelihood of HPV persistence and invasion and treatment relapses^5^. Furthermore, about 5% of all cervical cancer deaths are due to HIV.^2,6^ In turn, HPV infection increases the risk of HIV infection by approximately 2-2.5-fold.^7,8^ Their interconnected pathogenic mechanisms influence the progression of both HPV and HIV infections. HPV-induced immunosuppression enables the persistence of HPV, which in turn increases the risk of developing high-grade cervical intraepithelial lesions^9^.

Sierra Leone has an HIV prevalence of 1.7% among people aged 15-49 years, with over 60% of adults aged 15 and above living with HIV being women. ^10,11^ Data from our previous studies have shown enormous challenges with HIV care, including drug resistance mutations, high rates of late-stage HIV presentation, high in-hospital mortality, and high prevalence of co-infections^12–15^, portraying a severe immunocompromised state and high risk of cervical cancer^16^.

To address the burden of cervical cancer, the World Health Organization (WHO) launched the “90-70-90” targets under the Global strategy to accelerate the elimination of cervical cancer as a public health problem by 2030. This strategy proposes an increase in HPV vaccination among girls to 90% coverage, an increase in screening for cervical cancer to 70% by age 35 with a repeat at age 45, and 90% treatment coverage for women with pre-cancerous lesions and invasive cancer.^17^ While cervical cancer screening and management in women living with HIV have been incorporated into routine HIV services in many countries, Sierra Leone is yet to commence such services. There are no prior research data on HPV among women living with HIV in Sierra Leone. This study aimed to assess the prevalence and risk factors of HPV among women living with HIV at the national referral hospital in Sierra Leone. To the best of our knowledge, this is the first study evaluating HPV prevalence and genotype distribution among WLHIV in Sierra Leone.

## METHODS

### Study design and study population

We employed a cross-sectional study design to screen for HPV in women living with HIV aged 25 to 65, presenting at the Connaught Teaching Hospital’s HIV clinic for routine HIV services between September and December 2024.

### Study setting

We enrolled women living with HIV to collect primary data on cervical cancer at Connaught Hospital, which is Sierra Leone’s national referral hospital with a capacity of 300 beds. It is a teaching hospital affiliated with the College of Medicine and Allied Health Sciences of the University of Sierra Leone. Connaught Hospital’s HIV clinic is the largest HIV center in the country and doubles as the country’s only referral center for advanced HIV Diseases.

The HIV clinic offers outpatient services to over 7,500 clients as of December 2024. Out of 7,500, 4,567 (60.9%) were females, of whom 3,197 (42.6%) fell within the 25-65 year age range. The hospital offers inpatient services for HIV, which accounts for over 40% of all medical admissions^14^.

### Sample size

The primary aim of this study was to assess the prevalence of cervical dysplasia among WLHIV, for which the sample size was calculated using Fisher’s formula for single proportion studies (Z² p q/d²)^18^. A minimum sample size (n) of 178 was determined using a 95% confidence level (Z = 1.96), a cervical disease prevalence (p) of 0.134 reported in a comparable study conducted in Cambodia, a low- and middle-income country^19^, and its complement (q = 1.0 - p = 0.0.866) at a degree of precision (d) of 0.05. The first phase of this study is screening for HPV, the results of which have been prepared into this manuscript. Therefore, we used cervical dysplasia prevalence to calculate the sample size of this study.

### Sampling method

. Participants were selected using a systematic random sampling approach, whereby every fourth eligible woman who met the inclusion criteria and provided written informed consent was enrolled in the study.

### Inclusion and exclusion criteria

1. Women aged 25-65 years with confirmed HIV and no history of cervical diseases were eligible for inclusion if they were able and willing to provide written informed consent. We excluded pregnant women, women with a history of hysterectomy with removal of the cervix (trachelectomy) or prior diagnosis of cervical cancer or ongoing treatment for cervical cancer and women with active menstruation at the time of screening (with eligibility for rescheduling). We also excluded women with severe acute illness requiring immediate medical attention and those with an inability to provide informed consent due to cognitive impairment or severe illness.

### HPV DNA Screening and Testing

HPV DNA was determined using the High-Risk HPV Typing Nucleic Acid Detection kit (ZJ Biotech, Shanghai, China). The kit can detect 15 high-risk HPV types (16, 18, 31, 33, 35, 39, 45, 51, 52, 56, 58, 59, 66, 68, 82).

The high-risk HPV genotyping kits demonstrated a limit of detection (LoD) of 1×10⁴ copies/ml (defined via serial dilution of HPV-positive subtype standards). At this LoD, 10 independent repeated detections for each subtype yielded a 100% detection rate (no false negatives), meeting National Medical Products Administration (NMPA) In vitro Diagnostics registration Criteria. Analytical performance (LoD determination and reproducibility) was validated per the manufacturer’s protocol and NMPA guidelines.

● LoD Determination: Serial dilutions of HPV-positive subtype standards identified the lowest detectable concentration as 1×10⁴ copies/ml.
● Reproducibility: At this LoD, 10 independent repeated detections for each subtype showed a 100% detection rate (no false negatives; ZJ Bio-Tech validation report, 2024), confirming kit reliability for low-concentration samples.

Endocervical swabs were collected by trained clinicians according to standard protocols using a cytobrush and sent for HPV Deoxyribonucleic acid (DNA) testing.

Endocervical swab samples were processed for DNA extraction using the following protocol: Swabs were placed in 1.5 mL centrifuge tubes containing 1 mL of Lysis Buffer B and incubated at 56°C for 10 minutes. After incubation, 50 μL of Proteinase K was added, and the samples were further incubated at 56°C for 10 additional minutes. To precipitate DNA, 400 μL of absolute ethanol was added, followed by gentle mixing and a 5-minute incubation at room temperature. The mixture was then centrifuged at 13,000 rpm for 5 minutes, and the supernatant was carefully removed. The DNA pellet was washed with 1 mL of 70% ethanol, recentrifuged at 13,000 rpm for 5 minutes, and the supernatant was discarded. Each reaction mixture contained 36μL of HPV polymerase chain reaction (PCR) mixture 4μL of supernatant as DNA template. HPV amplification and detection were performed with the CFX96 real-time PCR detection system (Bio-Rad Laboratories, Hercules, California, USA).

### Data collection and analysis

A structured, interviewer-administered questionnaire was used to collect socio-demographic data (age, sex, occupation, level of education, history of cigarette smoking and illicit drug use, and marital status) and clinical data (parity, gravidity, use of contraceptives, abortion). HIV-related information was extracted directly from HIV registers and patients’ treatment cards. The questionnaire was developed in English and administered in the local Krio language to enhance participants’ understanding. Data collection was conducted by trained HIV nurses who were fluent in Krio and familiar with HIV and cervical cancer care protocols.

Statistical analyses were performed using R (R Core Team, Vienna, Austria). Baseline characteristics were summarized using frequency and percentage (%). We used Pearson’s chi-square and Fisher’s exact test to test statistical significance. Multivariable logistic regression was independently performed for two outcomes (HPV DNA positivity and multiple high-risk serotype co-infection), adjusting for age, education, occupation, marital status and parity. Candidate predictors underwent univariate screening (retaining variables with *P*<0.2), followed by multicollinearity assessment via variance inflation factors (VIF; sequentially removing variables with VIF>10 until all remaining had VIF≤10), final models were built using backward elimination of remaining variables, reporting adjusted odds ratios (aOR) with 95% confidence intervals (CI). The Hosmer-Lemeshow test indicated adequate fit (p = 0.78).

Statistical significance was considered with p-value < 0.05.

#### Variables

The primary outcome variable was high-risk HPV DNA positivity (binary: positive vs negative), defined as the detection of any of the 15 high-risk HPV genotypes by nucleic acid testing. The secondary outcome was multiple high-risk HPV co-infection, defined as detection of two or more high-risk HPV genotypes. Explanatory variables included socio-demographic characteristics (age, occupation, educational level, marital status), reproductive-health-related factors (age at first sexual intercourse, parity, age at first childbirth, abortion history and abortion times, contraceptive use status, contraceptive type and duration of contraceptive use), clinical HIV-related variables (HIV diagnosis duration, new patient status, antiretroviral treatment interruption), and behavioural factors (smoking history, alcohol drinking, illicit drug use). Family history of cancer and personal cancer history were also included as covariates. All categorical variables were coded according to pre-defined grouping criteria shown in Table 1.

**Table 1.**
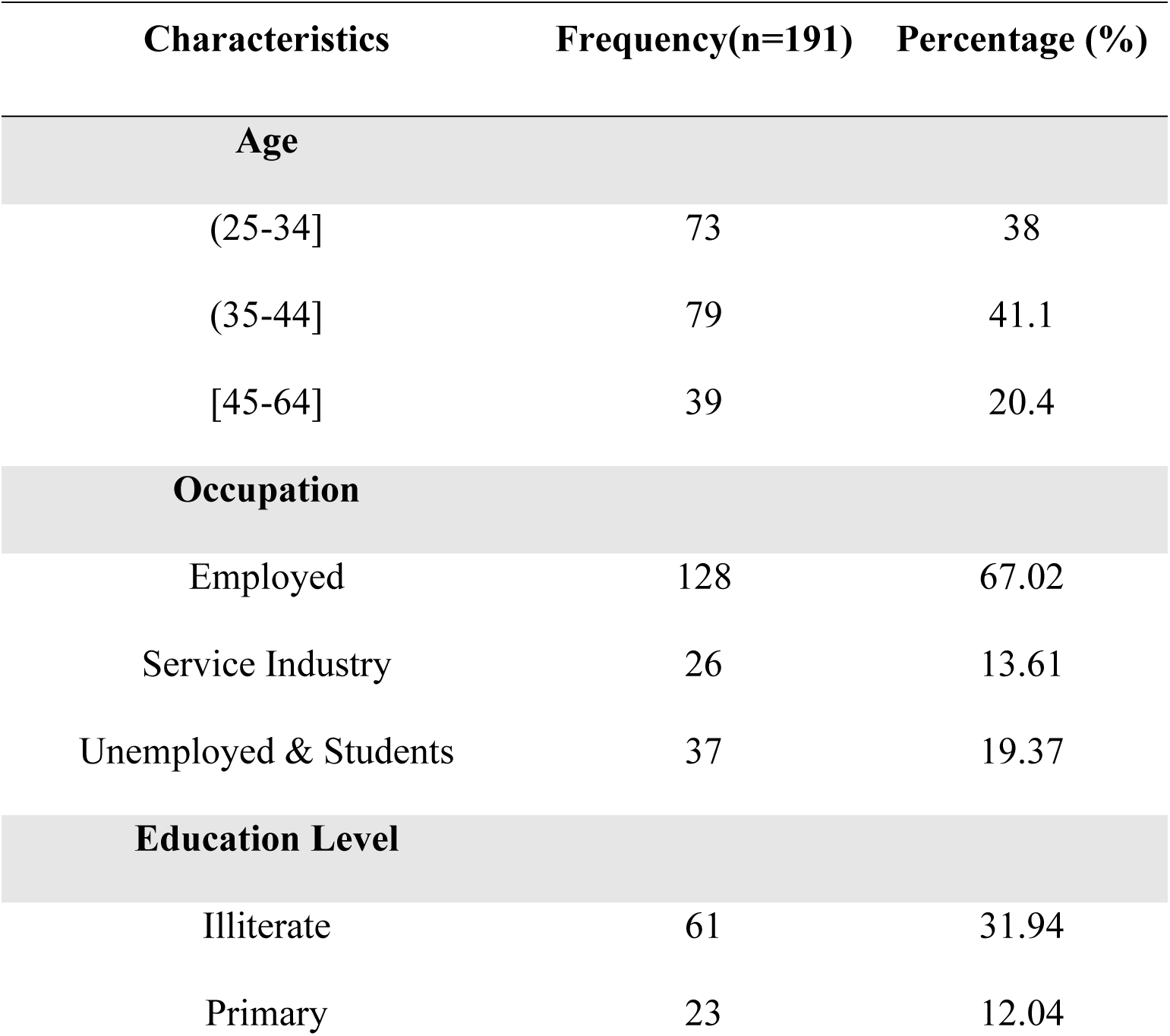
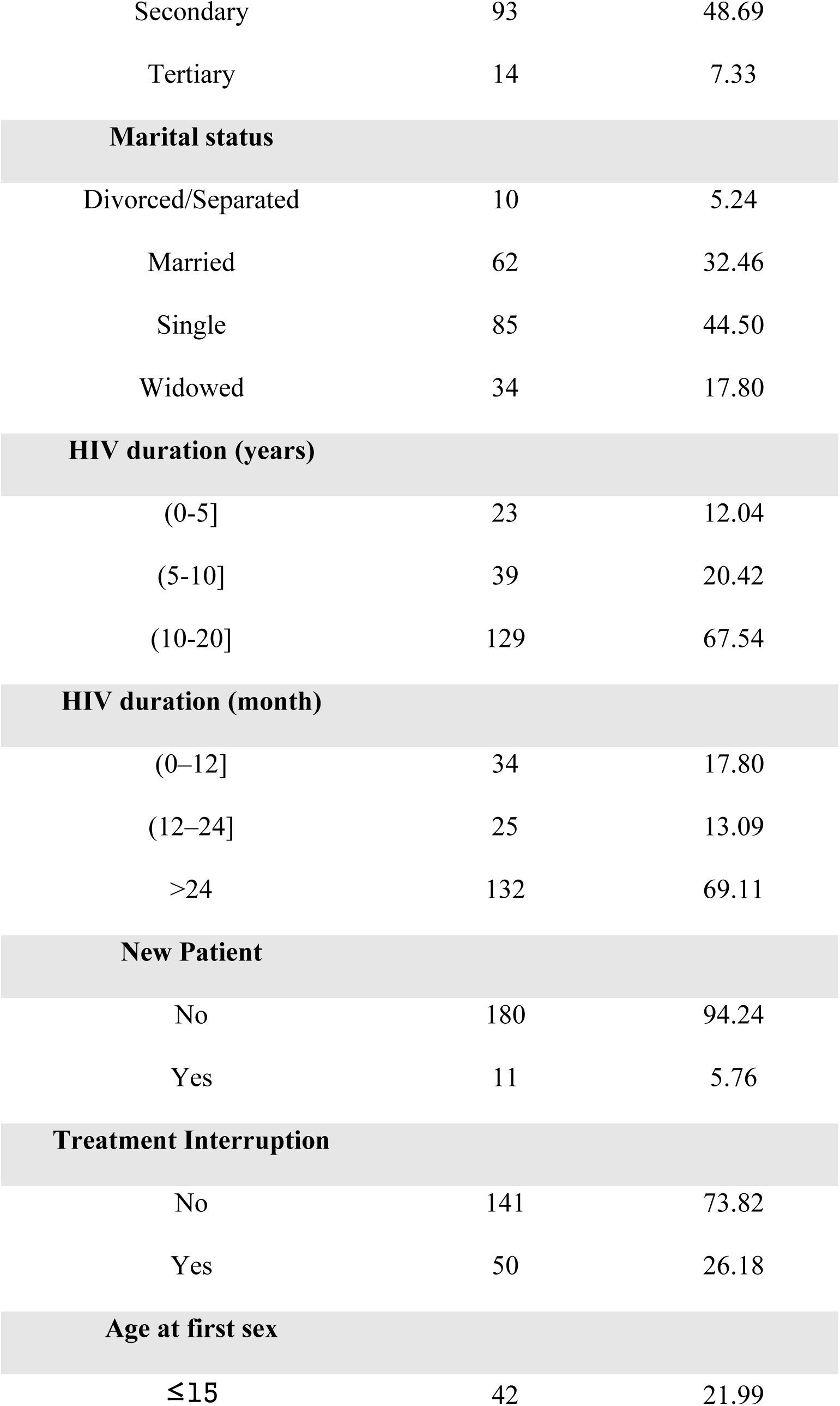
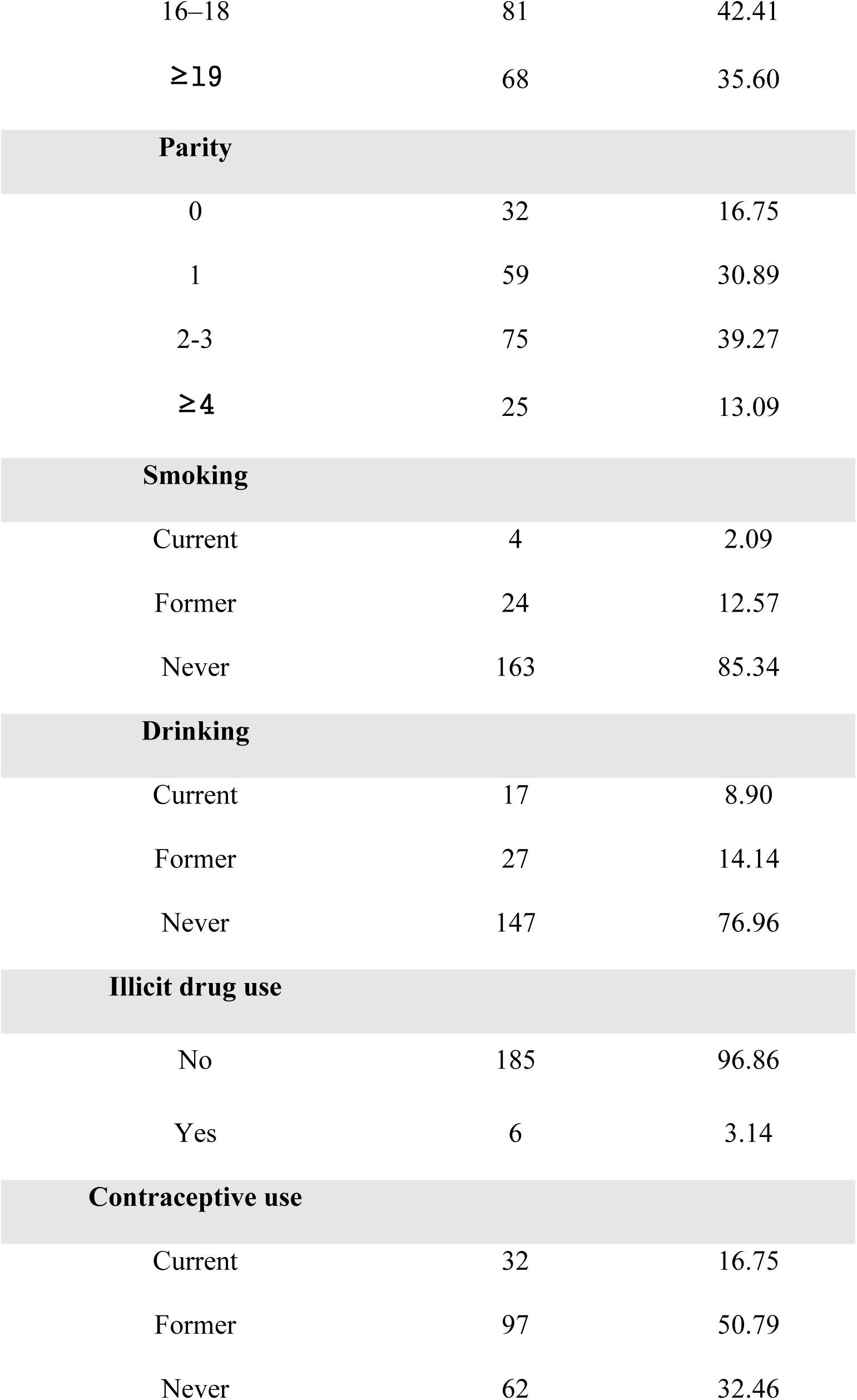
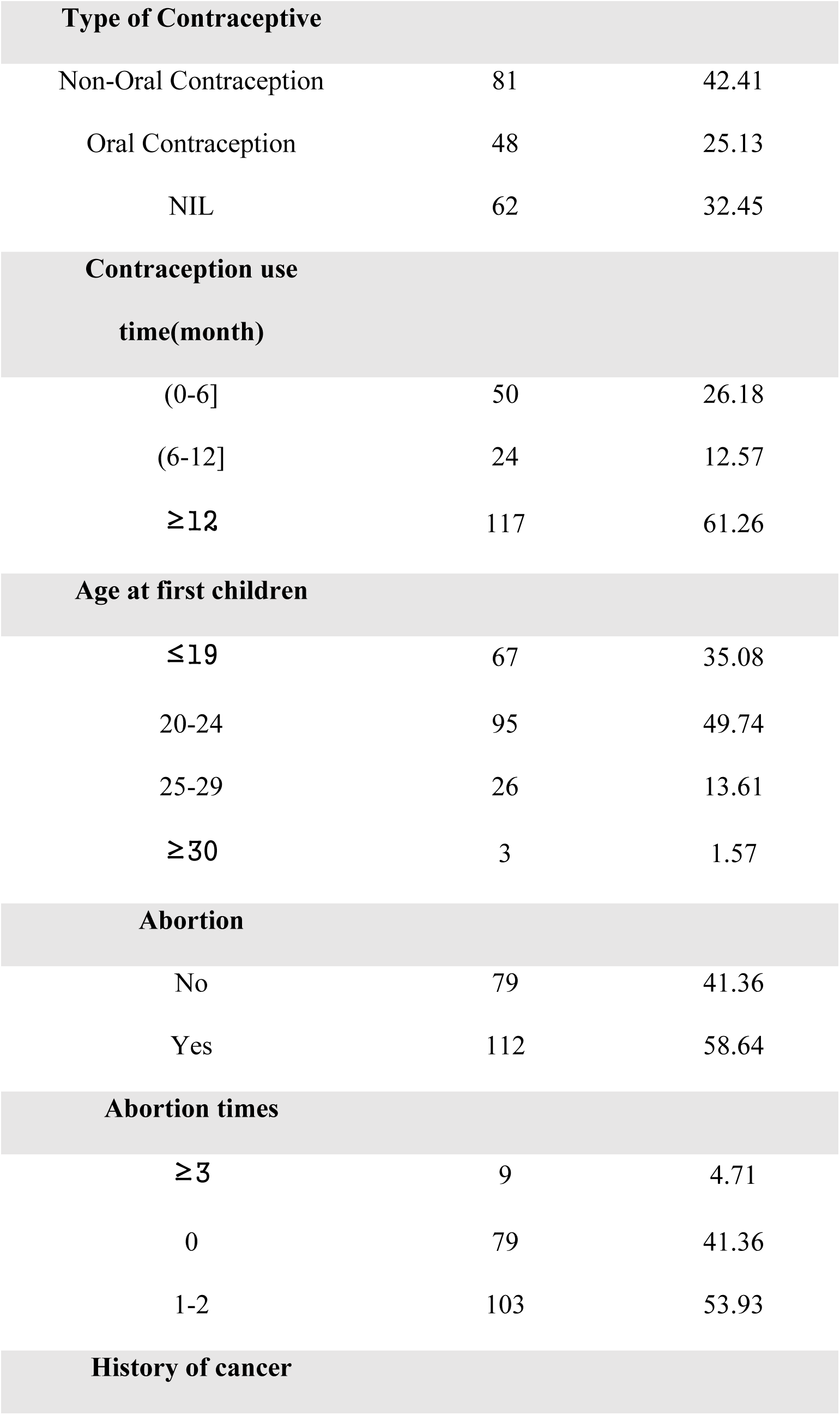
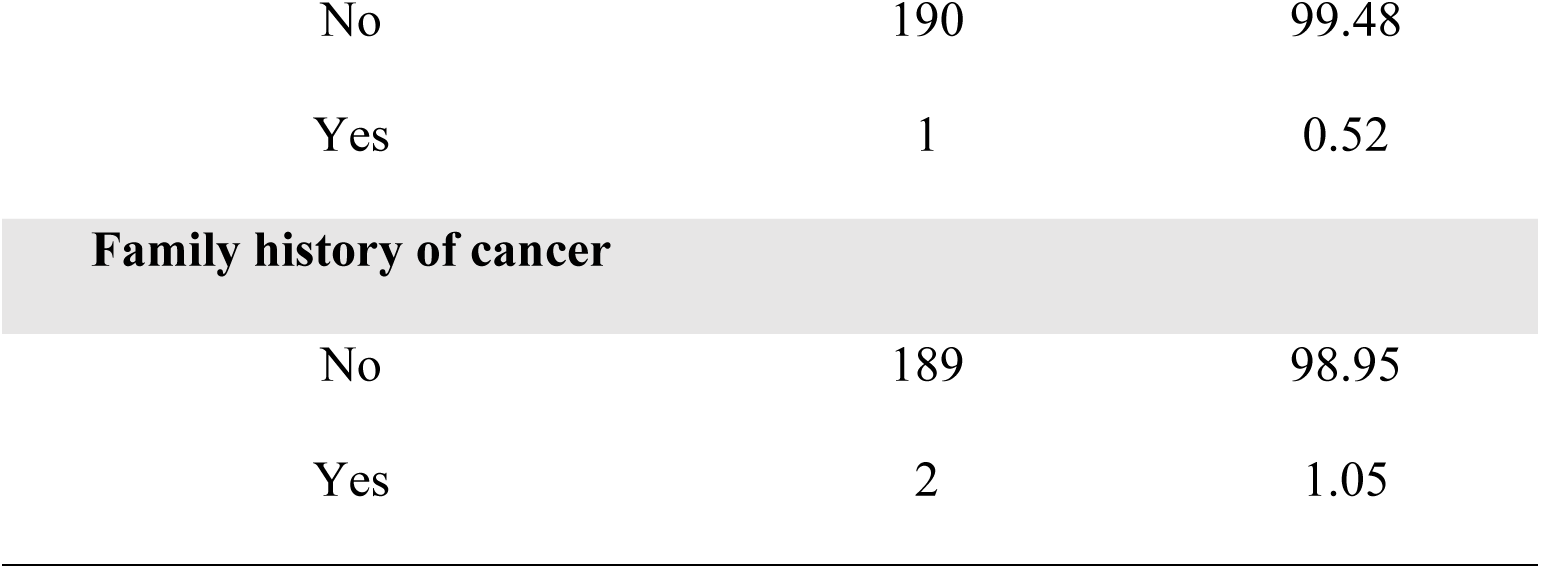
Socio-demographic and clinical characteristics of study participants (N = 191)

| Characteristics | Frequency(n=191) | Percentage (%) |
|---|---|---|
| **Age** |  |  |
| (25-34] | 73 | 38 |
| (35-44] | 79 | 41.1 |
| [45-64] | 39 | 20.4 |
| **Occupation** |  |  |
| Employed | 128 | 67.02 |
| Service Industry | 26 | 13.61 |
| Unemployed & Students | 37 | 19.37 |
| **Education Level** |  |  |
| Illiterate | 61 | 31.94 |
| Primary | 23 | 12.04 |
| Secondary | 93 | 48.69 |
| Tertiary | 14 | 7.33 |
| **Marital status** |  |  |
| Divorced/Separated | 10 | 5.24 |
| Married | 62 | 32.46 |
| Single | 85 | 44.50 |
| Widowed | 34 | 17.80 |
| **HIV duration (years)** |  |  |
| (0-5] | 23 | 12.04 |
| (5-10] | 39 | 20.42 |
| (10-20] | 129 | 67.54 |
| **HIV duration (month)** |  |  |
| (0–12] | 34 | 17.80 |
| (12–24] | 25 | 13.09 |
| >24 | 132 | 69.11 |
| **New Patient** |  |  |
| No | 180 | 94.24 |
| Yes | 11 | 5.76 |
| **Treatment Interruption** |  |  |
| No | 141 | 73.82 |
| Yes | 50 | 26.18 |
| **Age at first sex** |  |  |
| ≤15 | 42 | 21.99 |
| 16–18 | 81 | 42.41 |
| ≥19 | 68 | 35.60 |
| **Parity** |  |  |
| 0 | 32 | 16.75 |
| 1 | 59 | 30.89 |
| 2-3 | 75 | 39.27 |
| ≥4 | 25 | 13.09 |
| **Smoking** |  |  |
| Current | 4 | 2.09 |
| Former | 24 | 12.57 |
| Never | 163 | 85.34 |
| **Drinking** |  |  |
| Current | 17 | 8.90 |
| Former | 27 | 14.14 |
| Never | 147 | 76.96 |
| **Illicit drug use** |  |  |
| No | 185 | 96.86 |
| Yes | 6 | 3.14 |
| **Contraceptive use** |  |  |
| Current | 32 | 16.75 |
| Former | 97 | 50.79 |
| Never | 62 | 32.46 |
| **Type of Contraceptive** |  |  |
| Non-Oral Contraception | 81 | 42.41 |
| Oral Contraception | 48 | 25.13 |
| NIL | 62 | 32.45 |
| **Contraception use time(month)** |  |  |
| (0-6] | 50 | 26.18 |
| (6-12] | 24 | 12.57 |
| ≥12 | 117 | 61.26 |
| **Age at first children** |  |  |
| ≤19 | 67 | 35.08 |
| 20-24 | 95 | 49.74 |
| 25-29 | 26 | 13.61 |
| ≥30 | 3 | 1.57 |
| **Abortion** |  |  |
| No | 79 | 41.36 |
| Yes | 112 | 58.64 |
| **Abortion times** |  |  |
| ≥3 | 9 | 4.71 |
| 0 | 79 | 41.36 |
| 1-2 | 103 | 53.93 |
| **History of cancer** |  |  |
| No | 190 | 99.48 |
| Yes | 1 | 0.52 |
| **Family history of cancer** |  |  |
| No | 189 | 98.95 |
| Yes | 2 | 1.05 |

#### Data sources and measurement

Socio-demographic, behavioural, clinical and reproductive-health data were collected by trained research staff using standardized paper-based questionnaires administered through face-to-face participant interviews. Endocervical swab specimens were collected using cytobrushes for laboratory HPV-DNA testing. HPV genotyping was performed with the High-Risk HPV Typing Nucleic Acid Detection kit (ZJ Biotech, Shanghai, China), which could identify 15 high-risk HPV genotypes. DNA extraction and real-time PCR amplification were performed following the manufacturer’s standard laboratory protocols. Questionnaire variables and laboratory test results were double-entered and cleaned prior to statistical analysis.

#### Bias

Several potential sources of bias were addressed in this study. To reduce selection bias, we consecutively enrolled every fourth eligible female client attending the HIV clinic to avoid convenience-sampling bias; however, as recruitment was conducted within a single national referral hospital, selection bias could not be fully eliminated and the sample may not represent all women living with HIV across Sierra Leone. Interview-related information bias was minimised by training interviewers and using a standardised questionnaire. Laboratory measurement bias was reduced by strictly following standard operating procedures for specimen collection, DNA extraction and PCR detection. Missing data were minimal in this dataset. Confounding bias was managed in the analytical stage: univariate screening, variance inflation factor assessment for multicollinearity, and multivariable logistic regression with backward elimination were applied to adjust for potential confounding variables.

### Ethical Consideration

This study received ethical approval from the Sierra Leone Ethics and Scientific Review Committee of the Ministry of Health, Government of Sierra Leone (referenced SLESRC NO: 012/07/2023) in accordance with the relevant guidelines and regulations and the Declaration of Helsinki. All study participants provided written informed consent before enrollment and participation. Data were anonymized and entered into a password-protected computer accessible only to the study team.

## Results

We recruited 191 HIV-infected women with a modal age group of 25-34 years and a mean age of 34.61 years. Most were single (44.5%), employed (67%), and only 7% had a tertiary education. The mean duration of HIV diagnosis was 85 months (7 years). All participants were receiving antiretroviral therapy. The mean age at first sexual intercourse was 17.7 (±7.82 SD) years. About 22% had sex before 15 years, and only 32% had never used contraceptives in their lifetime (Table 1).

### Profile of HPV serotypes

Of the 191 women living with HIV, 60 (31%, 95% CI: 25.3% to 38.3%) tested positive for high-risk HPV, with half of them having more than one genotype. Genotype 52 was detected in 17 (28.3%) participants. Other detected genotypes were 16, 18, 31, 33, 35, 38, 38, 39, 45, 51, 55, 56, 58, 59, 68, and 82. Only 30% (n=18/60, 95% CI: 20-43%) had either genotype 16 or 18, with one participant having both 16 and 18 (Supplementary Table 1, Figure 1).

**Figure 1:**
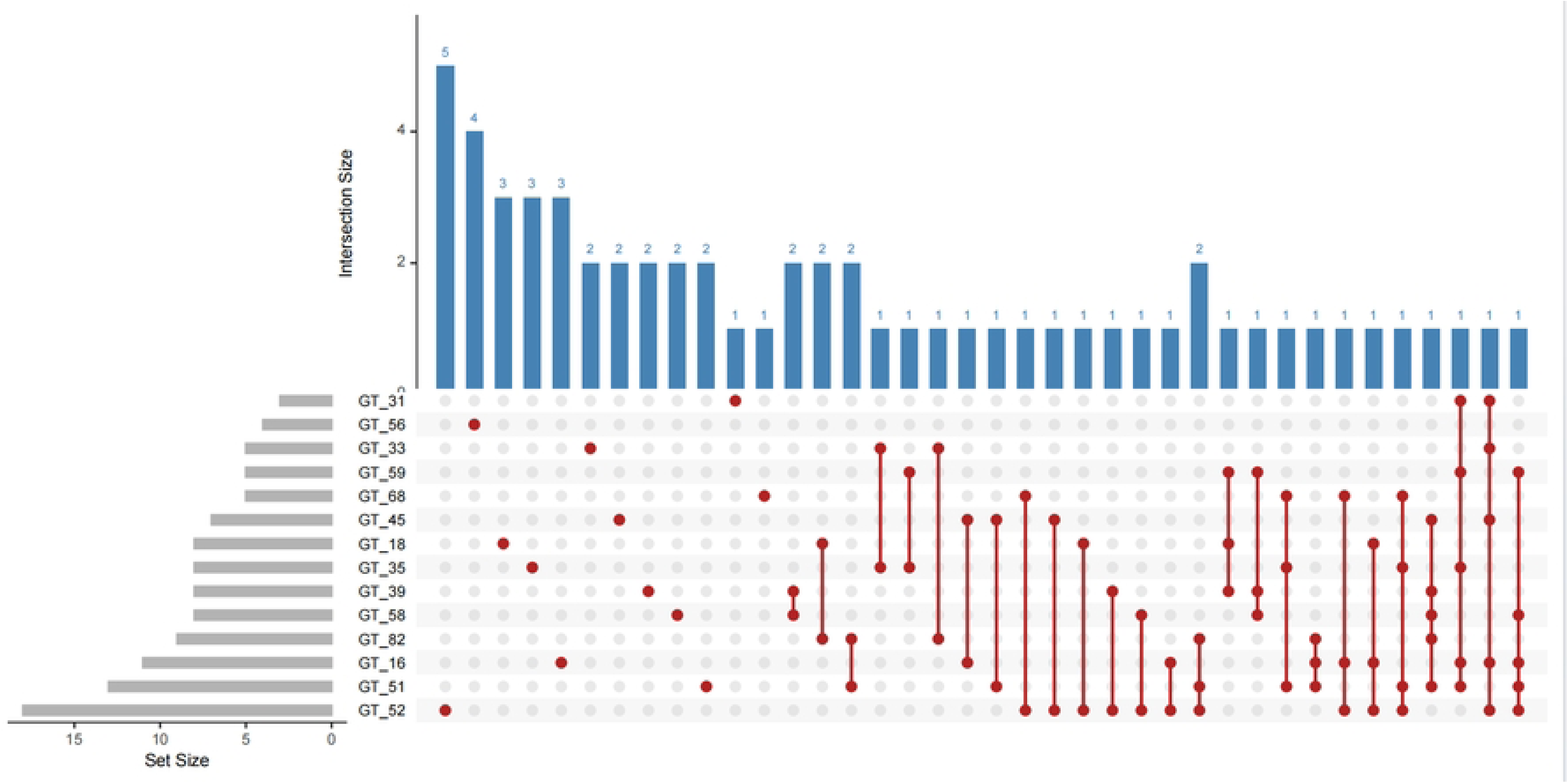
Distribution of HPV DNA serotypes among study participants. Red dots represent patients infected with a single serotype. Blue bars illustrate the total number of infections for each serotype, including both single infections and co-infections, while the grey bars indicate the overall number of patients affected by each serotype.

### Risk factors for HPV positivity

Current use of contraceptives is associated with HPV DNA positivity (X^2^; 6.87, p=0.032). Although not statistically significant, the HPV positivity rate was high in participants 25-34 years, single, employed, and those with secondary school education, and women who had never smoked cigarettes or drunk alcohol (Table 2).

**Table 2:**
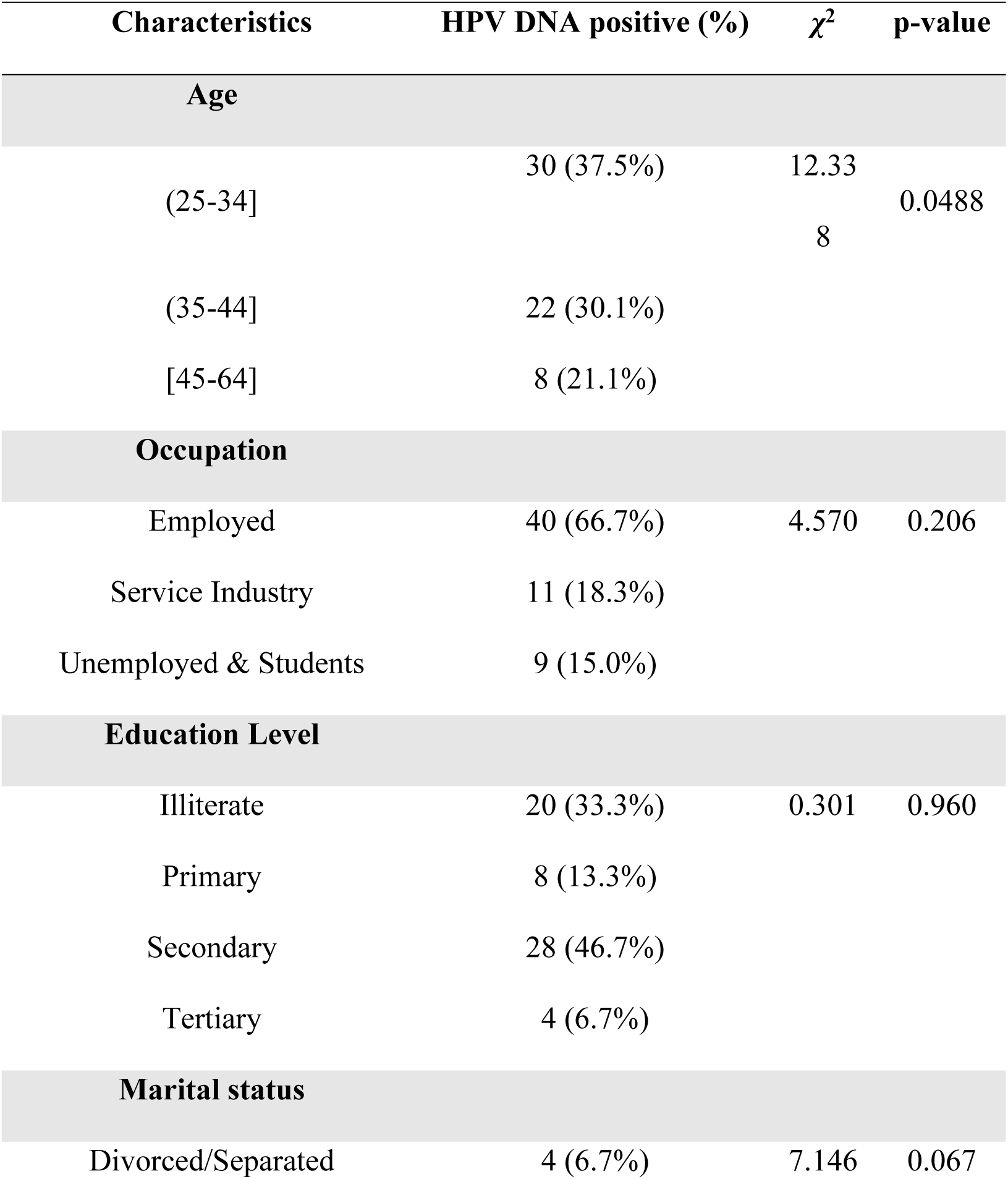
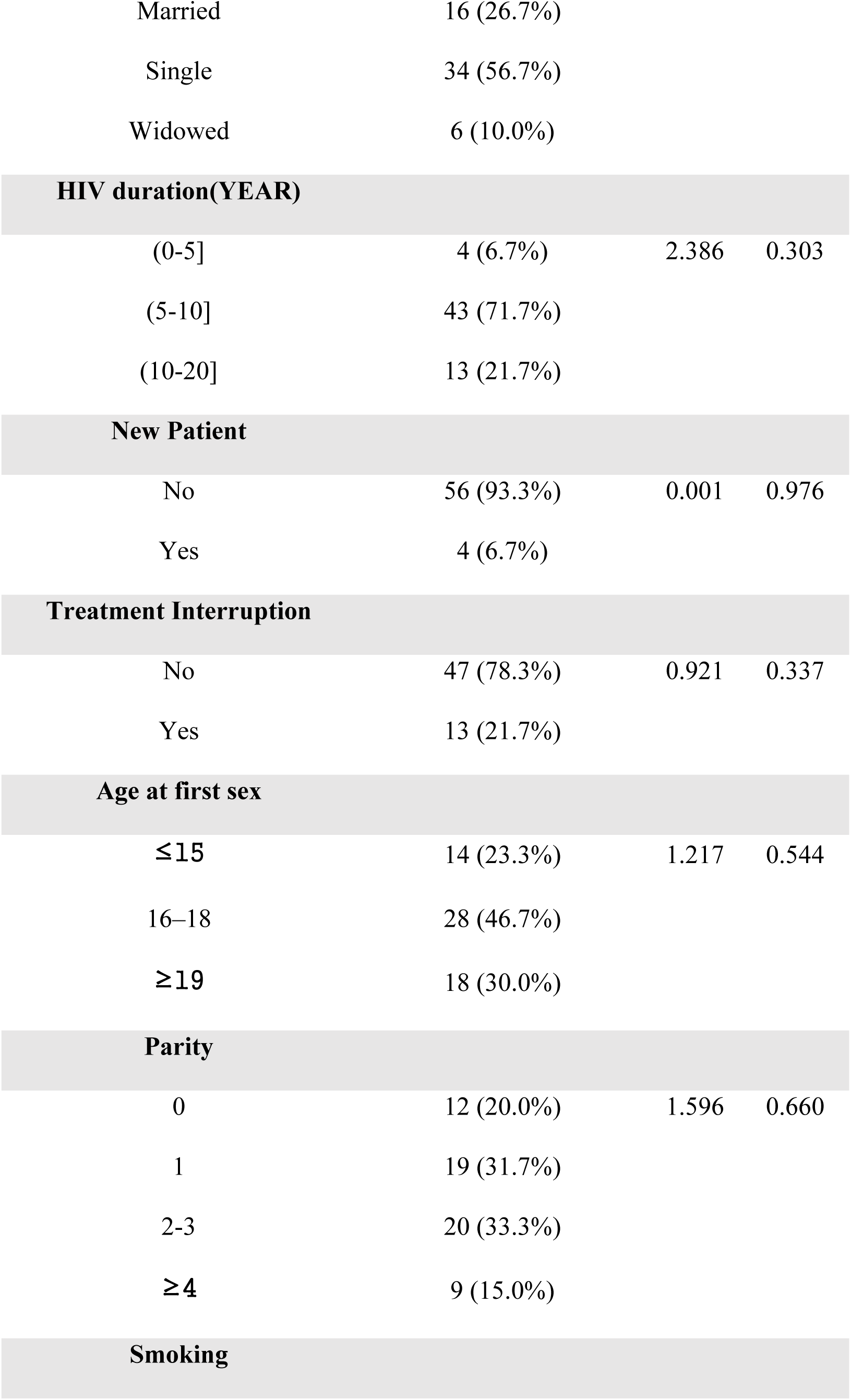
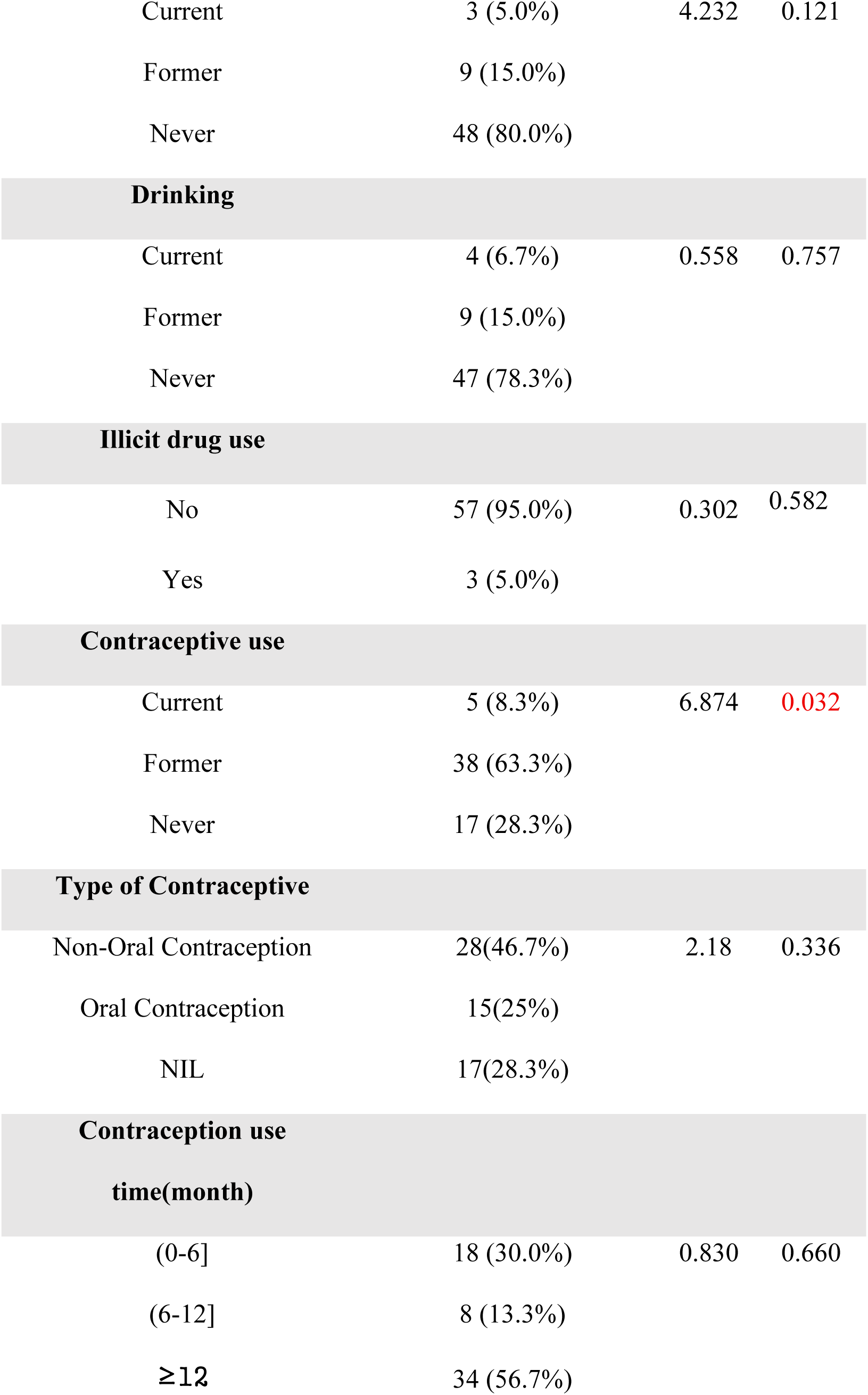
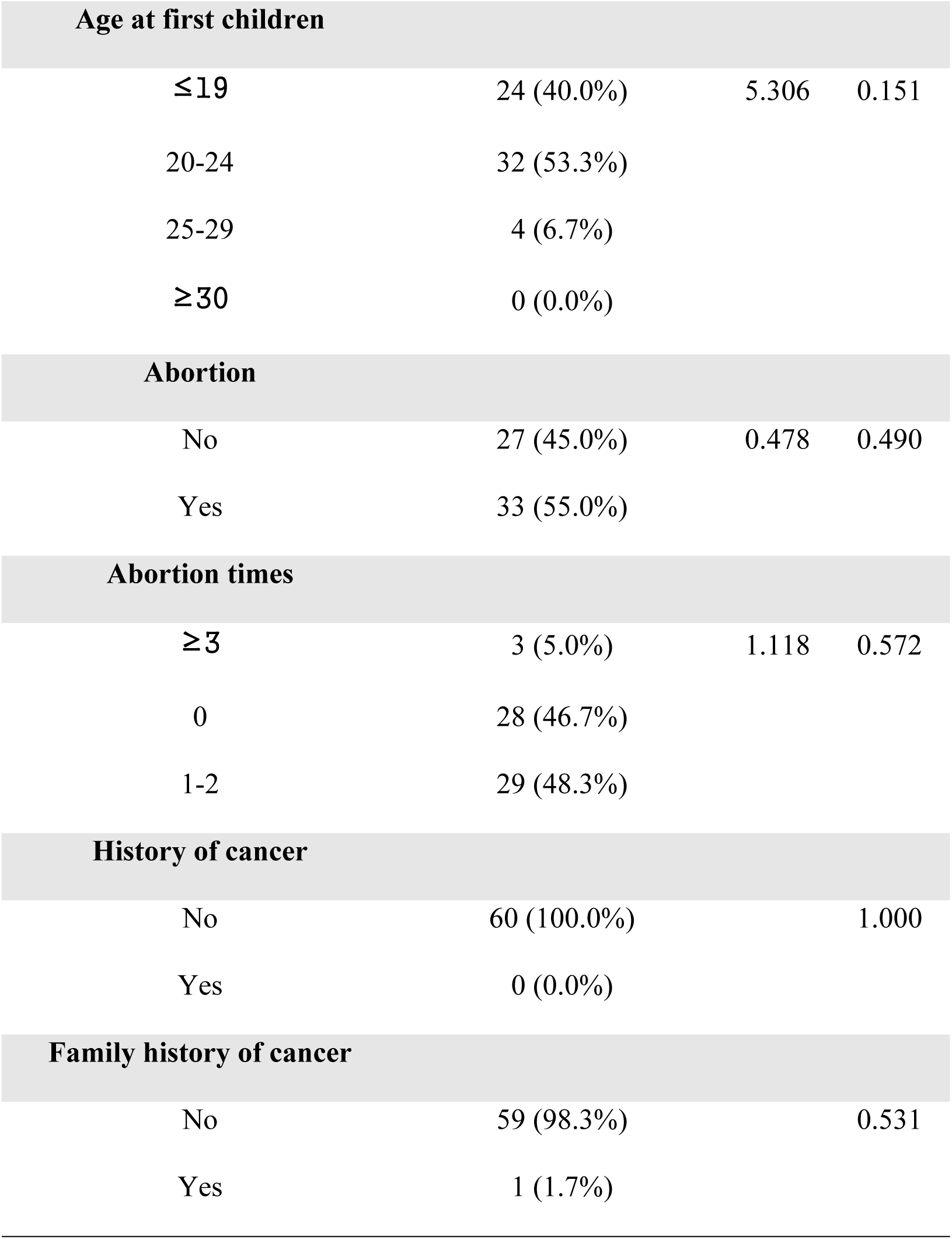
Relationship and distribution of studied risk factors to HPV DNA status.

| **Characteristics** | **HPV DNA positive (%)** | ***χ***<sup>**2**</sup> | **p-value** |
|---|---|---|---|
| **Age** |  |  |  |
| (25-34] | 30 (37.5%) | 12.338 | 0.0488 |
| (35-44] | 22 (30.1%) |  |  |
| [45-64] | 8 (21.1%) |  |  |
| **Occupation** |  |  |  |
| Employed | 40 (66.7%) | 4.570 | 0.206 |
| Service Industry | 11 (18.3%) |  |  |
| Unemployed & Students | 9 (15.0%) |  |  |
| **Education Level** |  |  |  |
| Illiterate | 20 (33.3%) | 0.301 | 0.960 |
| Primary | 8 (13.3%) |  |  |
| Secondary | 28 (46.7%) |  |  |
| Tertiary | 4 (6.7%) |  |  |
| **Marital status** |  |  |  |
| Divorced/Separated | 4 (6.7%) | 7.146 | 0.067 |
| Married | 16 (26.7%) |  |  |
| Single | 34 (56.7%) |  |  |
| Widowed | 6 (10.0%) |  |  |
| **HIV duration(YEAR)** |  |  |  |
| (0-5] | 4 (6.7%) | 2.386 | 0.303 |
| (5-10] | 43 (71.7%) |  |  |
| (10-20] | 13 (21.7%) |  |  |
| **New Patient** |  |  |  |
| No | 56 (93.3%) | 0.001 | 0.976 |
| Yes | 4 (6.7%) |  |  |
| **Treatment Interruption** |  |  |  |
| No | 47 (78.3%) | 0.921 | 0.337 |
| Yes | 13 (21.7%) |  |  |
| **Age at first sex** |  |  |  |
| ≤15 | 14 (23.3%) | 1.217 | 0.544 |
| 16–18 | 28 (46.7%) |  |  |
| ≥19 | 18 (30.0%) |  |  |
| **Parity** |  |  |  |
| 0 | 12 (20.0%) | 1.596 | 0.660 |
| 1 | 19 (31.7%) |  |  |
| 2-3 | 20 (33.3%) |  |  |
| ≥4 | 9 (15.0%) |  |  |
| **Smoking** |  |  |  |
| Current | 3 (5.0%) | 4.232 | 0.121 |
| Former | 9 (15.0%) |  |  |
| Never | 48 (80.0%) |  |  |
| **Drinking** |  |  |  |
| Current | 4 (6.7%) | 0.558 | 0.757 |
| Former | 9 (15.0%) |  |  |
| Never | 47 (78.3%) |  |  |
| **Illicit drug use** |  |  |  |
| No | 57 (95.0%) | 0.302 | 0.582 |
| Yes | 3 (5.0%) |  |  |
| **Contraceptive use** |  |  |  |
| Current | 5 (8.3%) | 6.874 | 0.032 |
| Former | 38 (63.3%) |  |  |
| Never | 17 (28.3%) |  |  |
| **Type of Contraceptive** |  |  |  |
| Non-Oral Contraception | 28(46.7%) | 2.18 | 0.336 |
| Oral Contraception | 15(25%) |  |  |
| NIL | 17(28.3%) |  |  |
| **Contraception use time(month)** |  |  |  |
| (0-6] | 18 (30.0%) | 0.830 | 0.660 |
| (6-12] | 8 (13.3%) |  |  |
| ≥12 | 34 (56.7%) |  |  |
| **Age at first children** |  |  |  |
| ≤19 | 24 (40.0%) | 5.306 | 0.151 |
| 20-24 | 32 (53.3%) |  |  |
| 25-29 | 4 (6.7%) |  |  |
| ≥30 | 0 (0.0%) |  |  |
| **Abortion** |  |  |  |
| No | 27 (45.0%) | 0.478 | 0.490 |
| Yes | 33 (55.0%) |  |  |
| **Abortion times** |  |  |  |
| ≥3 | 3 (5.0%) | 1.118 | 0.572 |
| 0 | 28 (46.7%) |  |  |
| 1-2 | 29 (48.3%) |  |  |
| **History of cancer** |  |  |  |
| No | 60 (100.0%) |  | 1.000 |
| Yes | 0 (0.0%) |  |  |
| **Family history of cancer** |  |  |  |
| No | 59 (98.3%) |  | 0.531 |
| Yes | 1 (1.7%) |  |  |

On multivariable analysis, former contraceptive use was independently associated with increased HPV positivity (aOR=4.86, 95% CI: 1.65-17.37, p = 0.0032) (Table 3). Similarly, former contraceptive use was an independent predictor of multiple HPV genotypes (aOR=4.6, 95% CI: 1.3-22.05, p=0.016) (Table 4).

**Table 3.** Multivariable logistic regression results for HPV DNA infection risk factors.

| Variables | aOR | 95% Cl | p-value |
|---|---|---|---|
| Marital status MARRIED | 0.47 | 0.1,2.19 | 0.3253 |
| Marital status SINGLE | 0.93 | 0.21,4.23 | 0.9237 |
| Marital status WIDOWED | 0.39 | 0.07,2.07 | 0.2654 |
| Age at first childbirth | 0.92 | 0.83,1 | 0.0583 |
| HIV duration(YEAR) | 0.96 | 0.89,1.03 | 0.2255 |
| Smoking FORMER | 0.47 | 0.03,5.1 | 0.5377 |
| Smoking NEVER | 0.27 | 0.02,2.49 | 0.2431 |
| Contraceptive use FORMER | 4.86 | 1.65,17.37 | 0.0032 |
| Contraceptive use NEVER | 3.33 | 0.94,13.53 | 0.0623 |

**Table 4.**
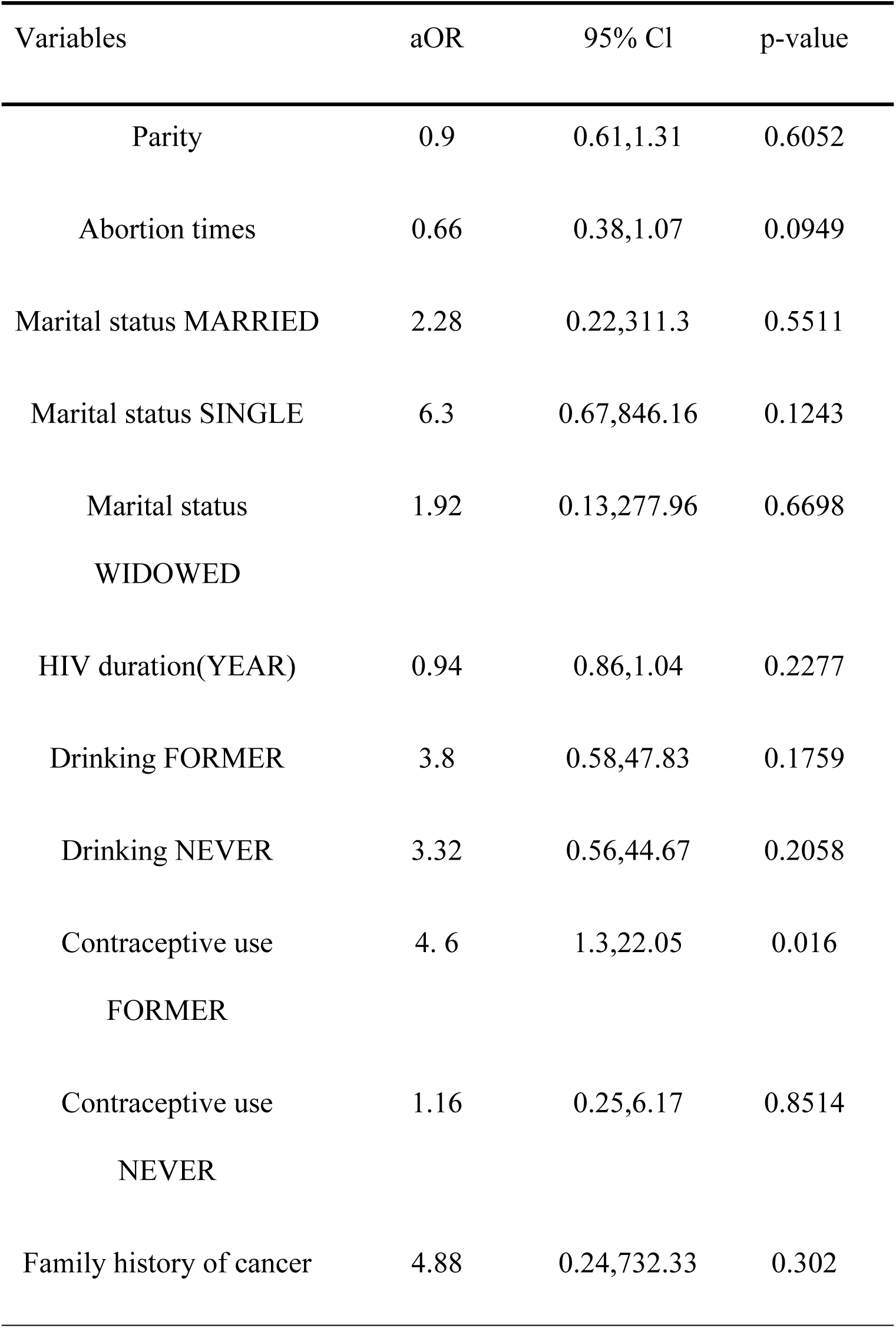
Multivariable logistic regression results for HPV multiple serotype infection risk factors.

## Discussion

This is the first study assessing the prevalence and risk factors of HPV infection among women living with HIV in Sierra Leone, providing insights on the burden of HPV infection. Our results report a 31% prevalence of high-risk HPV infections among women living with HIV with more than half of them having more than one genotype. Also, serotype 52 was detected in 17 (28.3%) participants. Only 30% of those who tested positive for HPV had either genotype 16 or 18, with one participant having both. Finally, former use of contraceptives is an independent predictor of HPV DNA positivity.

The high-risk HPV prevalence among women living with HIV (31.4%) from our study is similar to studies in women living with HIV in Europe (30.5-33.9%)^20^ and China (30.6%)^21^. Similar prevalence rates are reported among the general population in a systematic review in SSA (34%)^22^, and from observational studies in Nigeria (37%)^23^ and Ghana (37%)^24^. Furthermore, our high-risk HPV prevalence is higher than reported in Uganda (21%)^25^.

The high prevalence of high-risk HPV infection among women living with HIV in Sierra Leone reflects the country’s substantial burden of HIV and cervical cancer. Even though the HIV prevalence is 1.7% in Sierra Leone^4,11^, there is a high burden of advanced HIV disease^13,14,26^. Cervical cancer is the second most common cancer among women in Sierra Leone, with the country being placed at 22^nd^ in the world, for the estimated age-adjusted death rate^4^. There are limited cervical cancer services in the country, with screening and treatment services available in only 5 of the 16 districts. Even though the WHO recommends HPV testing as the first-choice method of screening for cervical cancer^17^, Sierra Leone still uses visual inspection with acetic acid in all 15 screening centres in the country^4^.

The main HPV genotypes reported in Eastern and Southern African countries are 16 and 52, whereas 16 and 35 are the main genotypes in Western African countries^27^. Unlike the reported genotypes across Africa, the commonest genotype in our study is 52, followed by 16 and 51. This is contrary to a recent systematic review in women living with HIV, which reported 16 as the most common genotype, followed by 18, and 45^28,29^. Our study reveals the high prevalence of multiple high-risk HPV infections (50%), higher than in the global report (39.1%) and studies from Africa (22.6%)^29,30^.

These multiple infections are known to play a significant role in cervical carcinogenesis due to increased persistence/prolonged exposure and high viral load to various types of high-risk HPVs, with possibilities of synergistic oncogenic effects^31–34^.

Reports have shown that smoking^35,36^, alcohol^37–39^, multiparity^40,41^, use of oral contraceptive pills^41–44^ and early age of sexual debut and first childbirth^45–47^ are some notable risk factors for persistent HPV infection and cervical cancer. These are significantly present in our participants, further solidifying their risk of persistent HPV infection and progression to cervical cancer, in the background of their already immunocompromised state. The lack of a statistically significant association between high-risk HPV genotypes and factors like smoking, alcohol use, and early sexual debut, contrary to other studies^35,37,41^, could be attributed to variations in sample size, methodological differences, or population characteristics.

Highly active antiretroviral therapy has been shown to reduce the incidence and progression of lesions in women living with HIV^48,49^. Even though all our participants in our study are on highly active antiretroviral therapy, 26.2% of them had treatment interruption, highlighting the need for continuous treatment adherence counseling for both new and old clients, given the benefits of sustained viral suppression.

Cervical cancer is preventable and highly treatable when detected early. Generally, cervical cancer screening in women living with HIV is low in SSA; (30%)^50^. The negligible uptake identified in our study is alarming, considering their significant risk of developing cervical cancer, as it represents a missed opportunity for early intervention, especially in a low resource setting like ours.

Our study reports that former use of oral contraceptives is an independent predictor of increased HPV positivity and the presence of multiple genotypes in women living with HIV. The use of hormonal contraceptives has been linked to oncogenesis and HPV persistence, likely due to hormonal effects on cervical endothelial changes, highlighting the importance of robust screening strategies for early detection and management of cervical cancer in women taking these contraceptives^51^.

Sierra Leone has piloted HPV vaccination in adolescent girls, with over 150,000 girls vaccinated. The vaccine used was a bivalent vaccine targeting HPV genotypes 16 and 18^4^. The 5 most common genotypes from our study were 52, 16, 51, 35, 39, and 58, justifying the need for quadrivalent or, better still, a nonvalent HPV vaccine with a high HPV genotype coverage.

Connaught Teaching Hospital’s HIV clinic, Sierra Leone’s national HIV referral centre, does not have structures for cervical cancer screening. As integration of cervical cancer services into routine HIV care has been shown to improve the quality of life of women living with HIV,^52^ we hope that these findings highlighted from our study will inform decisions and policies on implementation and integration of cervical cancer services in ART clinics in Sierra Leone.

Our study has limitations-it is a single-center study with a small sample size of 191 women living with HIV in a national referral hospital; thus, its findings are not representative of the HPV status in Sierra Leone. The high-risk HPV typing nucleic acid detection kit used in this study has a higher LoD of 1×10⁴ copies/ml. This may have underestimated the prevalence of high-risk HPV genotype in this population. Again, we did not measure CD4 cell count and assess its impact on HPV positivity. Despite this limitation, this study has clinical and public health relevance as it provides the first evidence of HPV in Sierra Leone.

## Conclusion

We report a high prevalence (31%) of high-risk HPV genotypes among women living with HIV in Sierra Leone, dominated by genotype 52, which is different from those protected against using the bivalent vaccines. The high prevalence of high-risk HPV and predominance of non 16/18 genotypes underscore the need for integrating HPV screening into HIV care, and introducing broader-spectrum vaccination (nonavalent vaccines) could substantially reduce cervical cancer risk among women living with HIV in Sierra Leone. Furthermore, we observed that former oral contraceptive use is an independent predictor of increased high-risk HPV positivity and the presence of multiple high-risk HPV genotypes, emphasizing the need for robust screening strategies for women on contraceptives.

## Data Availability

Data will be made available upon request to the corresponding author.

## List of Abbreviations

DNA: Deoxyribonucleic acid
HIV: Human Immunodeficiency Virus
HPV: Human papillomavirus
LMIC: Low- and Middle-Income Countries
PCR: Polymerase chain reaction
SSA: Sub-Saharan Africa
WHO: World Health Organization
WLHIV: Women living with Human Immunodeficiency Virus

## Availability of data and materials

Data will be made available upon request to the corresponding author.

## Conflicts of Interest

The authors declare no conflict of interest.

## Clinical trial number

not applicable.

## Funding

The study was self-funded by the authors.

## Authors’ contributions

DFJ, GY, and SL conceptualized and designed the research study. BD designed the sample collection procedure. SI, US and MASK collected samples from all participants. T.Z performed the laboratory tests. DFJ, ESN, UB, MNK, AK and DS collected the required data. LZ conducted data analysis and interpretation of the results. DF, GFD and OA drafted the initial version of the manuscript. All authors have reviewed multiple drafts, provided significant edits and feedback, and approved the final version for submission. SL, GY, JAD and BD oversaw the entire project, providing a critical review, guidance, and leadership throughout the research process.

## Acknowledgements

We would like to sincerely thank the management of Connaught Hospital for granting us the opportunity to conduct this study within their facility. We are also deeply grateful to the Directorate of Sexual and Reproductive Health, Ministry of Health, Sierra Leone for providing us with staff who supported the sample collection process. Their cooperation and commitment were invaluable to the successful completion of this research.

---

**Supplementary table S1:** HPV DNA serotypes among the study population.

| **HPV Serotype** | **n** | **% of HPV-positive (n=60)** | **% of total cohort (n=191)** |
|---|---|---|---|
| 16 | 11 | 18.3 | 5.8 |
| 18 | 6 | 10.0 | 3.1 |
| 31 | 3 | 5.0 | 1.6 |
| 33 | 4 | 6.7 | 2.1 |
| 35 | 4 | 6.7 | 2.1 |
| 39 | 6 | 10.0 | 3.1 |
| 45 | 4 | 6.7 | 2.1 |
| 51 | 5 | 8.3 | 2.6 |
| 52 | 17 | 28.3 | 8.9 |
| 56 | 4 | 6.7 | 2.1 |
| 58 | 3 | 5.0 | 1.6 |
| 59 | 2 | 3.3 | 1.0 |
| 68 | 2 | 3.3 | 1.0 |
| 82 | 4 | 6.7 | 2.1 |
| Multiple* | 30 | 50.0 | 15.7 |
| Negative | 131 | — | 68.6 |

\*Multiple：≥2 serotype
HPV 16-positive (HPV16+) means that HPV type 16 was detected, either alone or in combination with other HR types (e.g., 16 + 31, 16 + 52, etc.).
• HPV 18-positive (HPV18+) means that HPV type 18 was detected, either alone or in combination with other HR types (e.g., 18 + 45, 18 + 58, etc.).

## Notes

### Competing Interest Statement

The authors have declared no competing interest.

### Clinical Trial

NA

